# Seroprevalence of SARS-CoV-2 IgG antibodies, in Corsica (France), April and June 2020

**DOI:** 10.1101/2020.09.29.20201368

**Authors:** Capai Lisandru, Ayhan Nazli, Masse Shirley, Canarelli Jean, Priet Stéphane, Simeoni Marie Helene, Charrel Remi, de Lamballerie Xavier, Falchi Alessandra

## Abstract

Our aim was to assess the seroprevalence of severe acute respiratory syndrome coronavirus-2 (SARS-CoV-2) infection after the lockdown in a sample of the Corsican population. Between 16th April and 15th June 2020, 2,312 residual sera were collected from patients having carried out a blood analysis in one of the participating laboratories. Residual sera obtained from persons of all ages were tested for the presence of anti-SARS-CoV-2 IgG using the EUROIMMUN enzyme immunoassay kit for semiquantitative detection of IgG antibodies against S1 domain of viral spike protein (ELISA-S). Borderline and positive samples in ELISA-S were also tested with an in-house virus neutralization test (VNT). Prevalence values were adjusted for sex and age. A total of 1,973 residual sera samples were included in the study. The overall seroprevalence based on ELISA-S was 5.27% [95% confidence interval (CI) 4.33-6.35] and 5.46% [4.51-6.57] after adjustment. Gender was not associated with IgG detection. However, significant differences were observed between age groups (p-value = 1 E-5) and particularly for people being younger than 50 years of age (Odd ratio (OR) = 2.86 95% CI [1.80-4.53]; p-value <0.000001*). The prevalence of neutralizing antibody titers ≥40 was of 3% [2.28-3.84]. In conclusion the present study showed that a low seroprevalence for COVID-19 in Corsica in accordance with values reported for other French regions in which the impact of the pandemic was low.

## 1. Introduction

On December 30, 2019, the Municipal Health Commission in Wuhan (Hubei Province, China) reports a cluster of unexplained pneumonia cases (Ren *et al*., 2020). On January, 2020, a betacoronavirus, named the severe acute respiratory syndrome coronavirus (SARS-CoV-2) was identified (Chang *et al*., 2020; Zhou *et al*., 2020). The disease causes by the SARS-CoV-2 was named coronavirus infectious disease 2019 (COVID-19). COVID-19 is a highly infectious disease and following the first cases in China, the virus spread rapidly worldwide. Reasons for the rapid spread of SARS-CoV-2 include high transmissibility of the virus (Li *et al*., 2020a), asymptomatic or paucisymptomatic carriers (Bai *et al*., 2020), and the lack of any apparent cross-protective immunity from related viral infections (Loos *et al*., 2020). On January 30th, 2020 the World Health Organization (WHO) declared a Public Health Emergency of International Concern (Li *et al*., 2020b). As of September 4th, 2020, the number of SARS-CoV-2 confirmed cases exceeds 26 million and more than 800,000 deaths had been reported. The socioeconomic impact of the COVID-19 pandemic has also been significant, with lockdown drastically reducing the mobility and productivity of much of the world ‘s population worldwide (WHO, 2020).

In France, the SARS-CoV-2 epidemic period ranged from 2020w09(24-29 February) to 2020w19 (4-10 May), with an epidemic peak at 2020w13 (23-29 March) and a positivity rate of up to 30% affecting mostly the Paris region and the Northeastern of the country (Santé Publique France, 2020). To respond to the epidemic, on March 17th (2020w12), France ordered all non-essential retailers and services to be closed, and the general population to remain confined. These measures have reduced the number of incident cases and the stress on the health care system. The French government announced the end of the lockdown on May 11th (2020w21). After the epidemic period, it has been projected that in France, 3.7 million (range: 2.3–6.7) people, i.e., 5.7% of the population, will have been infected (Salje *et al*., 2020).

Corsica, a French Mediterranean island, was affected by the COVID-19 pandemic like continental France, at the beginning of March. The SARS-CoV-2, positivity rate ranged from 3.5% (2020w09) to 2.0% (2020w19), and peaked on 2020w13 at 14% (ARS, 2020). Until May 12th, 55 people died (46 in South Corsica, 9 in Haute-Corse) of COVID-19 in hospital (mortality rate = 0.0002% and lethality rate = 21.6%).

These epidemiological data included only a fraction of the real number of SARS-CoV-2 infections, since not all infected patients were symptomatic, required hospitalizations, or provided specimens for laboratory testing (Mizumoto *et al*., 2020). The capacity to estimate the spread of SARS-CoV-2 depends on our knowledge of the immune status against SARS-CoV-2 in the population.

The primary outcome of this study was to estimate for the first time the prevalence of IgG antibodies against SARS-CoV-2 in Corsican population in order to improve epidemiological knowledge of the virus spread and to estimate what part of the Corsican population has been infected by the SARS-CoV-2. The secondary outcome was to estimate neutralizing antibodies against SARS-CoV-2 by using an in-house virus neutralization test (VNT) currently considered as the most specific and sensitive serological assay capable of evaluating and detecting, functional neutralizing antibodies. Monitoring of seroprevalence can guide public health measures to fight the pandemic.

### 2. Experimental Section

### 2.1 Study area and population

The study was conducted in the French Mediterranean island of Corsica (8,680 km2) located south-east of mainland France. The population of the island was estimated at 344,679 as of January 1, 2019 (INSEE, 2020). This region is composed of two administrative departments (Haute-Corse and Corse-du-Sud) and five districts (Ajaccio, Bastia, Calvi, Corte, Sartène) including 365 counties. The age and sex distribution by age groups of the Corsican population was obtained from the French national institute of statistics and economic studies (INSEE, 2020) (Appendix 1; Fig 1A).

**Figure 1.**
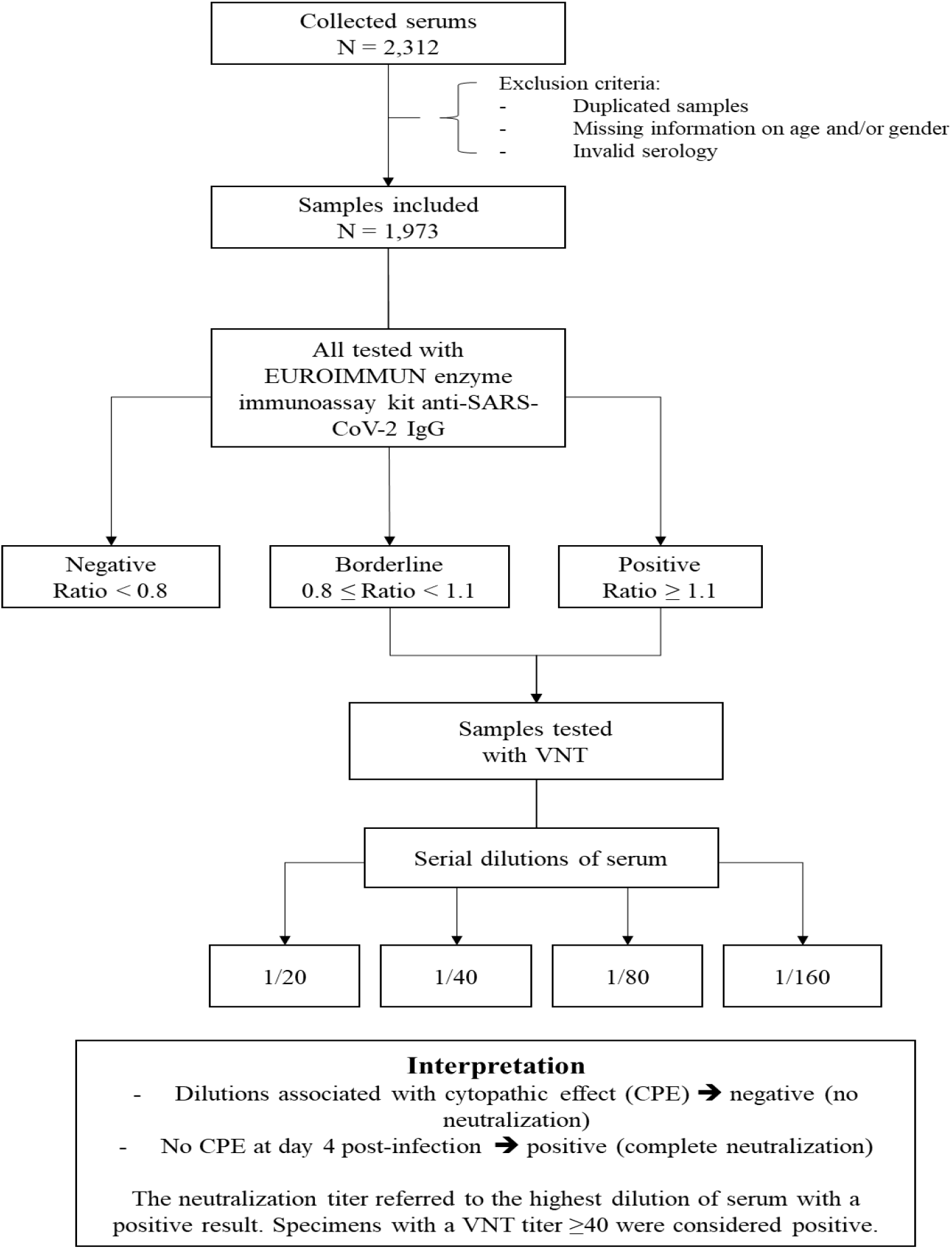
Diagram on sample inclusion and analyses performed.

### 2.2 Sample size

The sample size was calculated according to Epidemiological Calculation Tools (Epitools, 2018). A minimum sample size of 1814 was calculated assuming an a priori 5% IgG anti-SARS-CoV-2 seroprevalence (Salje *et al*., 2020), a confidence in the estimate of 95%, a maximum allowable error in the prevalence of 1%, and a Corsican population size of 344,679 habitants based on the latest French census data (INSEE, 2020).

### 2.3 Studied population

The sampling plan was established taking into account the minimum estimated sample size and the actual distribution of the Corsican population by age group and sex. Information about the distribution of the studied population and the general population of Corsica by age group and sex is available in the Appendix (Figure A1A and A1B).

Between 16th April and 15thJune 2020, 2,312 residual sera were collected from patients having carried out a blood analysis in one of the two participating laboratories (Figure 1). The conditions of exclusion were the following: one or more samples from the same person (only one sample included in the study), sample with missing information on age and/or gender and/or invalid serology result (insufficient serum volume or invalid result). The two private participating laboratories are located in Ajaccio and Corte. Ajaccio is one of the two largest cities of Corsica (south-west) and corresponds to the area of the island where the vast majority of COVID-19 cases were detected during the SARS-CoV-2 epidemic. The laboratory located in Ajaccio, is one of the main laboratory of the island and analyze samples coming from the south and north-western of the island. The laboratory of Corte analyzes samples from the center of Corsica.

### 2.4 Serological analyses

Residual sera obtained from persons of all ages were tested for the presence of anti-SARS-CoV-2 IgG using the EUROIMMUN enzyme immunoassay kit for semiquantitative detection of IgG antibodies against S1 domain of viral spike protein (ELISA-S) (reference: EI 2606-9601 G; EUROIMMUN, Bussy-Saint Martin, France). Wells are coated with recombinant structural protein of SARS-CoV-2. For each sample, the optical density (OD) ratio was estimated. According the manufacturer ‘s instruction, a result was considered borderline if the ratio was ≥ 0.8 and <1.1 and positive if the sample ratio was ≥ 1.1.

In all samples with a ratio ≥ 0.8, neutralizing antibodies were detected using a VNT as previously described (Gallian *et al*., 2020). VeroE6 cells cultured in 96-well microplates, 100 TCID50 of the SARS-CoV-2 strain BavPat1 (courtesy of Pr. Drosten, Berlin) and serial dilutions of serum (1/20–1/160) were used. Dilutions associated with cytopathic effect (CPE) were considered as negative (no neutralization) and those with no CPE at day 4 post-infection were considered positive (complete neutralization), respectively. The neutralization titer referred to the highest dilution of serum with a positive result. Specimens with a VNT titer ≥40 were considered positive (Gallian *et al*., 2020) (Figure 1).

### 2.5 Ethical statement

No nominative or sensitive data on participant people have been collected. This seroprevalence study falls within the scope of the French Reference Methodology MR-004 according to 2016–41 law dated 26 January 2016 on the modernization of the French health system. University Of Corsica Pascal Paoli (IRB UCPP 2020-01) approved this study.

### 2.6 Collection data and statistical analysis

According to French Reference Methodology MR-004 regulation on research projects concerning tube bottoms, only the age, sex, date and laboratory place of collection could be collected for each sample. The statistical analyses have been realized for these variables.

Descriptive statistic methods were performed for age and sex. Age was described as median with interquartile ranges (IQRs). Age groups were categorized as follow: 0-9; 10-19; 20-29; 30-39; 40-49; 50-59; 60-69; 70-79; 80-89 and ≥ 90. All categorical data were reported as percentages. IgG anti-SARS-CoV-2 seroprevalences and its 95% exact binomial confidence intervals (CIs) were estimated. Associations of the presence of anti-SARS-CoV-2 IgG with sex and age and location were analyzed and tested using the χ2 test or Fisher ‘s exact test. Statistical significance was set at a p value < 0.05. The odds ratio (ORs) was used to describe the risk of different gae groups and gender in positive ELISA-S serums compared with non-positive ELISA-S serums. Seroprevalence by age group and sex will be adjusted according to the proportions observed in the real population (INSEE, 2020). This adjustment will be made by specifically weighting each individual. The following R packages were used for the statistical analysis: questionr; car; stats; survey; FactoMineR and ade4. All statistical analyses were performed using R software version 3.6.1 (R Foundation, Vienna, Austria).

## 3. Results

A total of 1,973 serums of patients were included from the two participating private medical laboratories between late April and June 2020. People included ranged in age from 6 months to 101 years including 1,187 women (60.2%) and 786 men (39.8%). The median and the mean age were 52 years (interquartile 34-70). The weighted population according to sex and age groups, showed that the mean age was 44 years, including 51.7% of women and 48.3% of men.

### 3.1 Seroprevalence estimated with results of the EUROIMMUN ELISA IgG anti-SARS-CoV-2

The Table 1 and Figure 2 described the overall seroprevalence and the seroprevalence rates estimated by age groups and gender in the included and in the weighted population. One hundred four of 1,973 collected samples were found positive for anti-SARS-CoV-2 (index ≥1.1). The overall seroprevalence in our study population was of 5.27% [4.33-6.35] and of 5.46% [4.51-6.57] after adjustment. The median age among the positives was 41 years and the mean age was 44.92 years. Gender was not associated with seroprevalence rate. In contrast, univariate analysis showed significant seroprevalence rate differences by age groups (p-value = 1 E-5). The highest rate was observed in adults aged 40-49 years (10.48%) followed by 10-19 years (10.44%) and 30-39 years (8.88%). The seroprevalence values among the 0-9 years old and 20-29 were significantly lower with respect to the seroprevalence reported in 10-19, 30-39 and 40-49 age groups (p= 0.004 and 0.03, respectively). Individuals being aged less than 50 years old had a seroprevalence rate significantly higher (7.60%) than people aged more than 50 years old (2.80%) (OR = 2.86; 95% confidence intervals (CIs):1.80-4.53; p <0.000001).

**Table 1.**
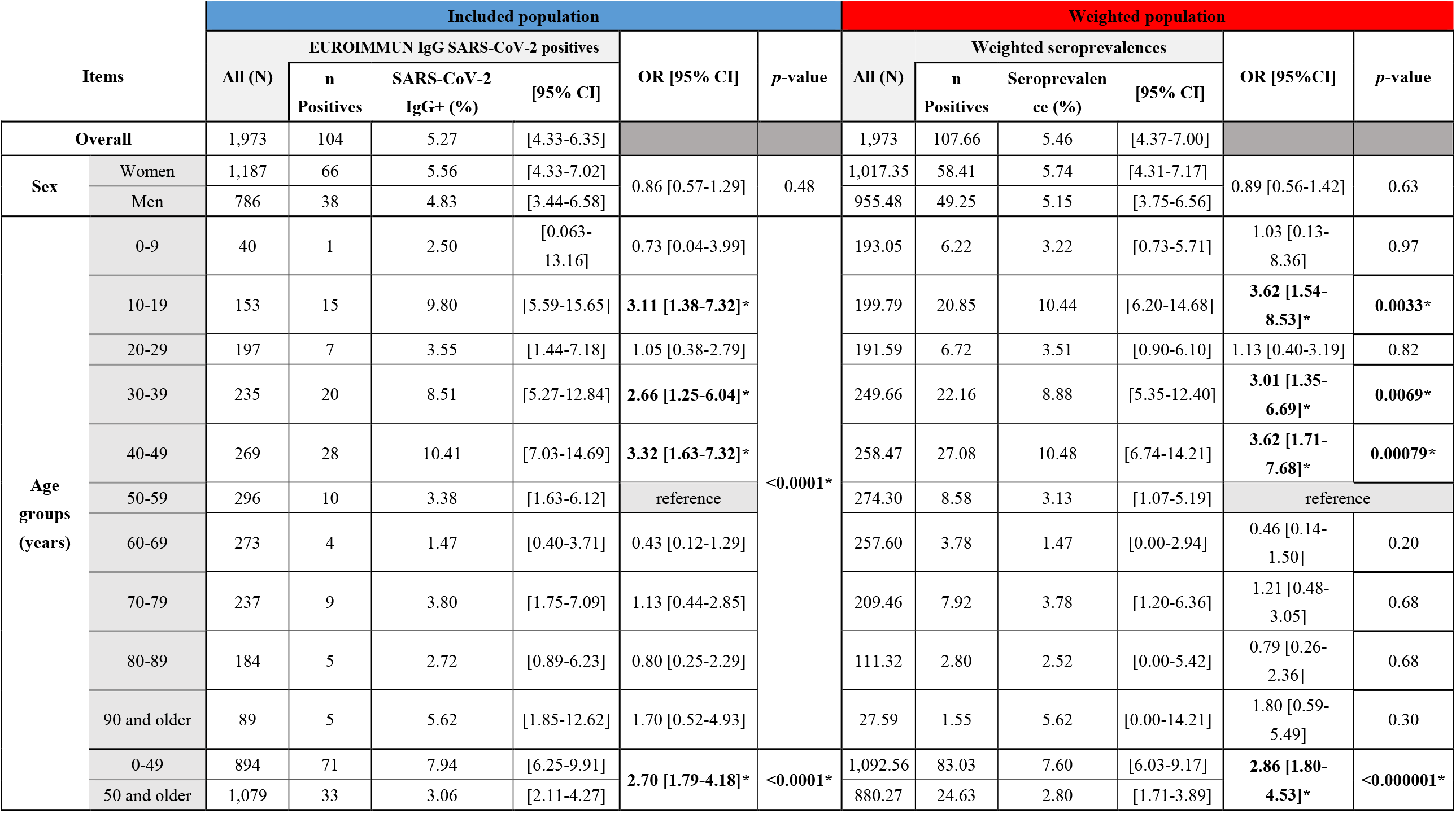
Description of the population (included and adjusted) and univariate analysis of association with IgG anti-SARS-CoV-2 seropositivity (p-value and Odd ratio).

**Figure 2.**
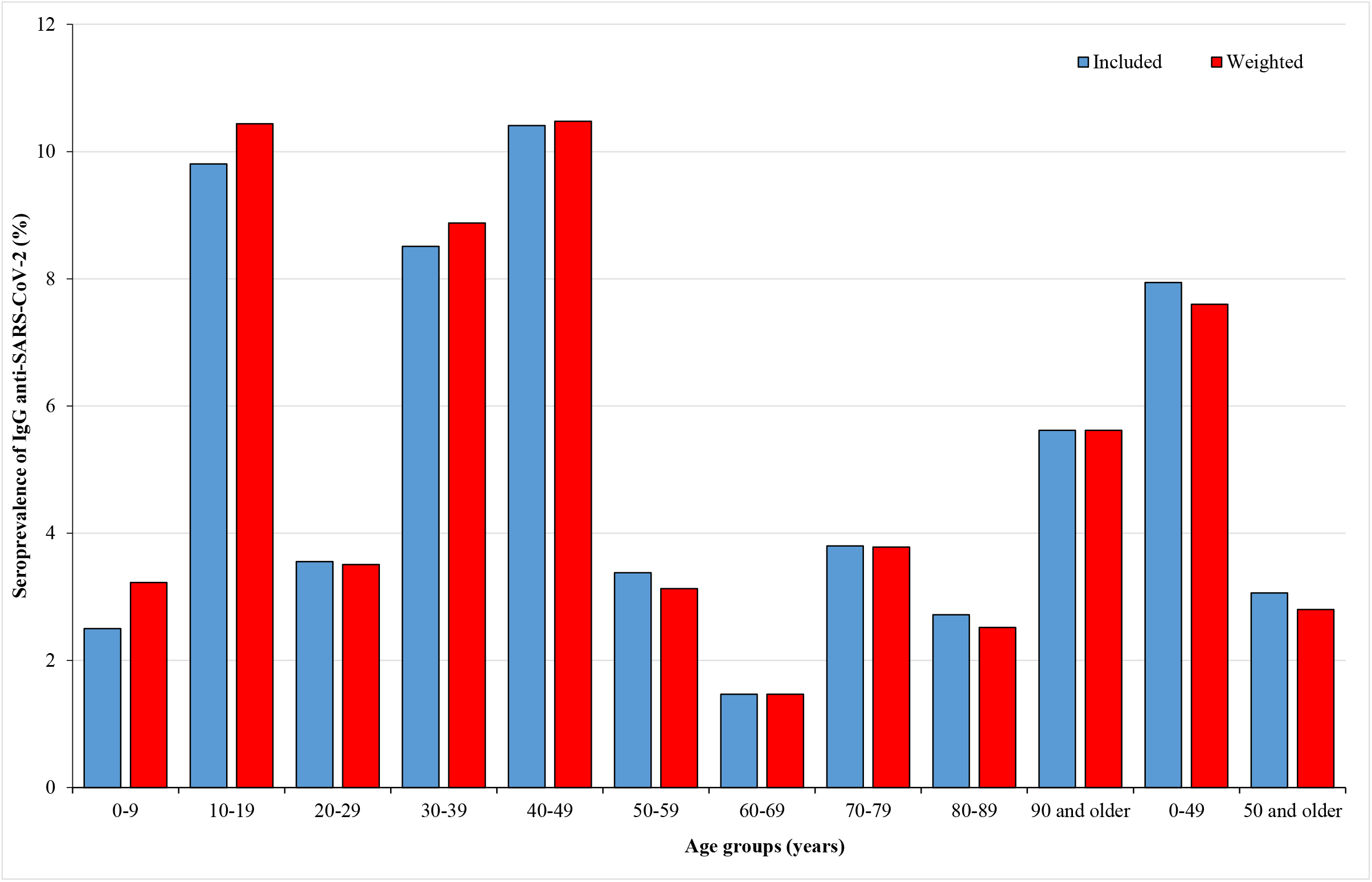
Seroprevalences of IgG anti-SARS-CoV-2 by age groups (included and weighted population).

### 3.2 Results of VNT assay

Among the 140 samples with an ELISA ratio ≥ 0.80 (Table 2), 104 samples showed a ratio >1.1 and 36 a ratio ranging from 0.8 to 1.1. Forty-two percent (n=59) of 140 samples had a positive neutralization antibody titer (VNT titer ≥ 40) (Table 2).

**Table 2.**
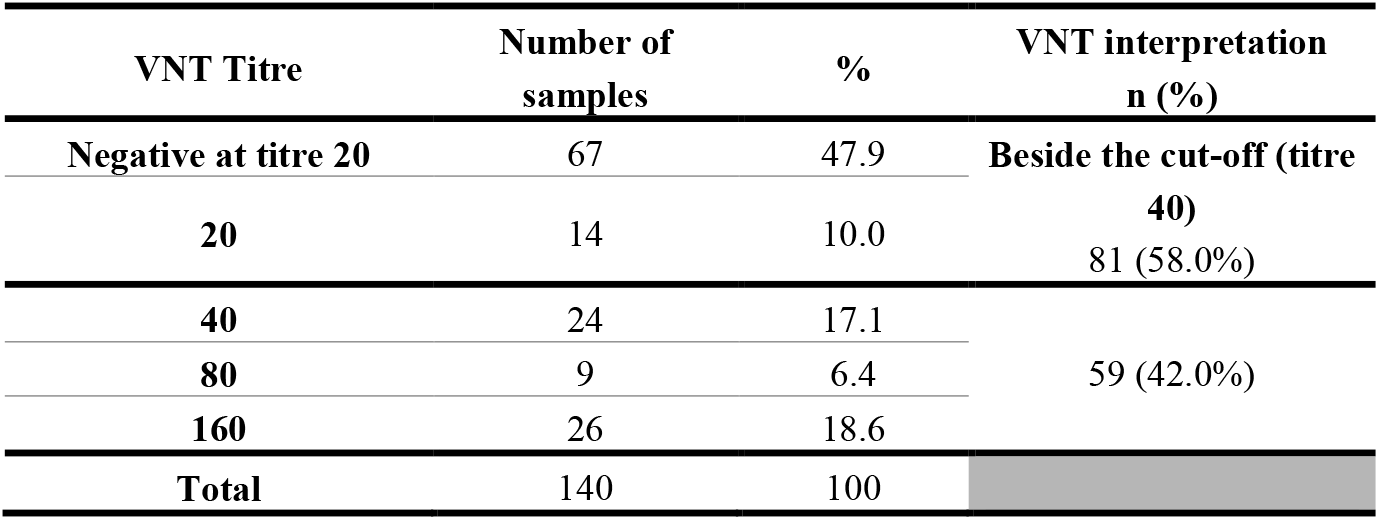
Virus neutralization test (VNT) results of 140 samples with an ELISA ratio >0.8.

| VNT Titre | Number of samples | % | VNT interpretation<br>n (%) |
| --- | --- | --- | --- |
| Negative at titre 20 | 67 | 47.9 | Beside the cut-off (titre 40)<br>81 (58.0%) |
| 20 | 14 | 10.0 |  |
| 40 | 24 | 17.1 | 59 (42.0%) |
| 80 | 9 | 6.4 |  |
| 160 | 26 | 18.6 |  |
| Total | 140 | 100 |  |

Among the 104 ELISA with a ratio>1.1, 53.8% (n=56) had a VNT titre ≥40. Among the 36 samples with an ELISA ratio between 0.80 and 1.1, 91.6% (n=33) had a VNT titre <40 and 8.3% (n=3) had a positive VNT titre ≥40. VNT titers did not differ significantly among age groups and similar VNT values were observed between males et females. The overall prevalence of samples above the cut-off (titre 40) (59/1973) was of 3% [2.28-3.84].

## 4. Discussion

To the best of our knowledge this is the first study describing the prevalence of SARS-CoV-2 antibodies in a representative sample of Corsican patients having carried out a blood analysis in biological laboratories after the COVID19 epidemic period.

The seroprevalence value estimated in the present study with ELISA-S (5.46% [4.51-6.57]; approximately 18,800 of people) was in line with an estimation that 3.7 million (range: 2.3–6.7) people, i.e., 5.7% of the French population, will have been infected during the epidemic period (Salje *et al*., 2020). The rate of seroprevalence reported in Corsica was closer to the rate reported by a similar study in a French region with low proportion of COVID-19 cases during the epidemic period (3%), than those reported in two regions with the highest rates (9-10%) (Carrat *et al*., 2020).The observed seroprevalence in our population was in line with ELISA-S values reported in Spain, New York city and in different subcohorts of Wuhan (Pollan *et al*., 2020), United States (Havers *et al*., 2020) and China (Xu *et al*., 2020) but lower than values reported in heavily affected areas such as Swiss (Stringhini *et al*., 2020), Northern Italy (Vena *et al*., 2020) and the urban areas around Madrid (Soriano *et al*., 2020).

In the present study we did not observed a significantly distribution of seroprevalence values between males and females, in agreement with previous reports in Spain (Pollan *et al*., 2020), Dutch blood donors (Slot, 2020) and French blood donors (Gallian *et al*., 2020). This is in line with sex-disaggregated data for COVID-19 in several European countries showing a similar number of cases between the sexes, but more severe outcomes in aged men (Gebhard *et al*., 2020). We observed that seroprevalence values differed significantly among age groups. Individuals aged less than 50 years old showed a seroprevalence rate significantly higher than people aged more than 50 years old. This trend was explained by high seroprevalence values observed among 10-19; 30-39 and 40-49 age groups ranging around 8-10%. This is in line with results previous reported by a population-based serosurvey in Geneva (Stringhini *et al*., 2020) and with the values reported in three French general populations (Carrat *et al*., 2020). In our sample seroprevalence values suggest that infection was less prevalent in children than in adolescents during the epidemic period. The lower prevalence in children might be in part related to lower nasal gene expression of angiotensin-converting enzyme 2 (Bunyavanich *et al*., 2020).

Because it is possible that the ELISA assay could exhibit cross-reactivity with antibodies to other seasonal human coronaviruses, some results may represent false-positives leading to overestimation of seroprevalence data (Takahashi *et al*., 2020). The EUROIMMUN assay used in this study was evaluated in different studies showing a specificity ranging from 96.2% to 100% and sensitivity ranging from 86.4% to 100% (Beavis *et al*., 2020; Kohmer *et al*., 2020 ; Kruttgen *et al*., 2020). The sensitivity and specificity of the ELISA can vary considerably depending on the timing of the sample, in particular from samples collected ≥4 days, after COVID-19 diagnosis by PCR. The sensitivity can increase from 67% to 100% between samples analyzed at the time of PCR and them analyzed at least four days later (Beavis *et al*., 2020). Virus neutralization test, currently considered as the gold standard, is the most specific serological assay capable of detecting true positive cases and functional neutralizing antibodies (Algaissi A, 2020). The positivity of a titer ≥40 for the VNT assay used in the present study is an indicator of strong specificity (100%) (Gallian *et al*., 2020). Around half of the samples showing ELISA results ≥0.8 half showed a VNT≥40 in line with a previous study (Carrat *et al*., 2020). The prevalence value based on our VNT analysis revealed that at least 3% of the included samples have been exposed to the virus and had developed a titer of neutralizing antibodies ≥40. This value is in line with the low value of 2.7% observed with the same VNT assay in blood donors collected during the last week of March or the first week of April 2020 in four French departments (Gallian *et al*., 2020) and in the adult general population of a region with low prevalence (Carrat *et al*., 2020).

Seroprevalence data based on ELISA assay provided information about previous exposure to SARS-CoV-2 but not about protection. In the present study we also estimated SARS-CoV-2 neutralizing antibodies titers among serum samples with dubious (≥0.8 and <1.1) and positive (≥ 1.1) results. About half of these samples had neutralizing antibodies with a titer ≥40. This represents a preliminary approach to protection against re-infection, but it should be underlined that no precise correlate of protection is available yet (Wu, 2020).

There are several limitations in our study. Firstly, residual sera from screening or routine care provided by private medical biology laboratories, are more likely to come from people needing to monitor their health status for chronic medical conditions. Thus data cannot be extrapolated to the general population although we have adjusted the data according to the age and sex of the general Corsican population. Additionally, no data concerning clinical features, chronic disease, or possible COVID-19 exposures were available potentially biasing results. Moreover, this lack of information on the COVID-19 status of the persons included could also influence the specificity and sensitivity of the ELISA test (timing of the sample in relation to the infection). As we tested by seroneutralization samples with an ELISA-S test optical density ≥0.8 and not all samples, seoprevalence could be slightly underestimated.

The strengths of this study were the size of the sample and its representativity in terms of age and gender. Samples have been analysed by combining ELISA and neutralization methods to strengthen results.

In conclusion the present study showed that a low seroprevalence for COVID-19 in Corsica in accordance with values reported for other French regions in which the impact of the pandemic was low. This regional study is particularly important in Corsica, as the island situation cannot be extrapolated from neighboring regions.

## Data Availability

No nominative or sensitive data on participant people have been collected. This seroprevalence study falls within the scope of the French Reference Methodology MR-004 according to 2016 41 law dated 26 January 2016 on the modernization of the French health system.

## Author Contributions

Conceptualization: A.F., M-H.S., J.C., R.C. and L.C.; Investigation: M-H.S. and J.C.; Funding acquisition: A.F.; methodology: R.C., A.F. and X.d.L, writing original draft preparation: L.C. and A.F.; Writing-review and editing: X.d.L., R.C. and A.F.; Laboratory analyses: S.M., N.A., S.P. and L.C.; Statistical analyses: L.C.; Supervision: A.F., M-H.S., J.C., R.C. and X.d.L.

## Appendix

**Figure A1.**
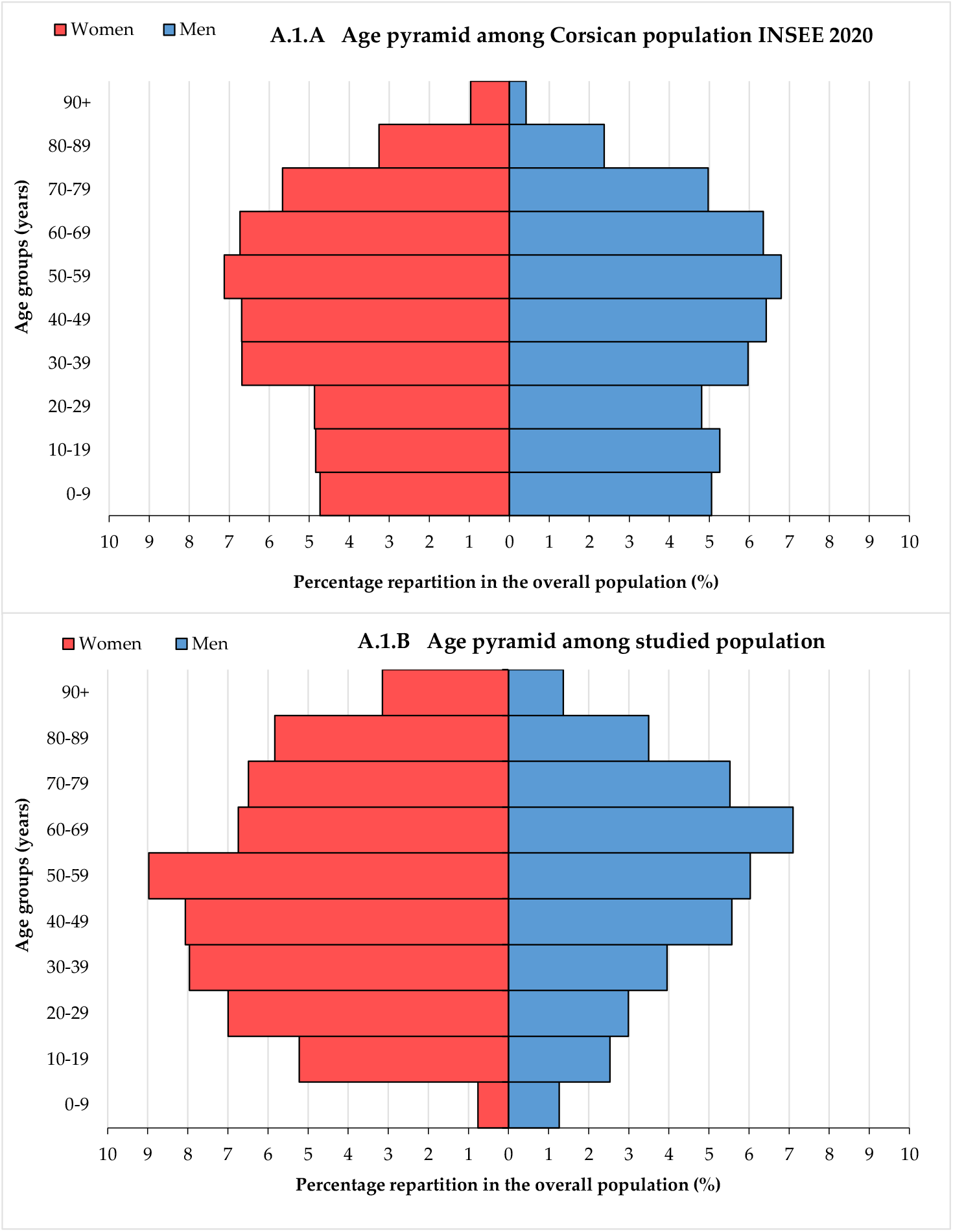
Age and sex repartition pyramid among Corsican population (A1A) and studied population (A1B).

